# Risk prediction for poor outcome and death in hospital in-patients with COVID-19: derivation in Wuhan, China and external validation in London, UK

**DOI:** 10.1101/2020.04.28.20082222

**Authors:** Huayu Zhang, Ting Shi, Xiaodong Wu, Xin Zhang, Kun Wang, Daniel Bean, Richard Dobson, James T Teo, Jiaxing Sun, Pei Zhao, Chenghong Li, Kevin Dhaliwal, Honghan Wu, Qiang Li, Bruce Guthrie

## Abstract

**Background:** Accurate risk prediction of clinical outcome would usefully inform clinical decisions and intervention targeting in COVID-19. The aim of this study was to derive and validate risk prediction models for poor outcome and death in adult inpatients with COVID-19.

**Methods:** Model derivation using data from Wuhan, China used logistic regression with death and poor outcome (death or severe disease) as outcomes. Predictors were demographic, comorbidity, symptom and laboratory test variables. The best performing models were externally validated in data from London, UK.

**Findings:** 4.3% of the derivation cohort (n=775) died and 9.7% had a poor outcome, compared to 34.1% and 42.9% of the validation cohort (n=226). In derivation, prediction models based on age, sex, neutrophil count, lymphocyte count, platelet count, C-reactive protein and creatinine had excellent discrimination (death c-index=0.91, poor outcome c-index=0.88), with good-to-excellent calibration. Using two cut-offs to define low, high and very-high risk groups, derivation patients were stratified in groups with observed death rates of 0.34%, 15.0% and 28.3% and poor outcome rates 0.63%, 8.9% and 58.5%. External validation discrimination was good (c-index death=0.74, poor outcome=0.72) as was calibration. However, observed rates of death were 16.5%, 42.9% and 58.4% and poor outcome 26.3%, 28.4% and 64.8% in predicted low, high and very-high risk groups.

**Interpretation:** Our prediction model using demography and routinely-available laboratory tests performed very well in internal validation in the lower-risk derivation population, but less well in the much higher-risk external validation population. Further external validation is needed. Collaboration to create larger derivation datasets, and to rapidly externally validate all proposed prediction models in a range of populations is needed, before routine implementation of any risk prediction tool in clinical care.

**Funding:** MRC, Wellcome Trust, HDR-UK, LifeArc, participating hospitals, NNSFC, National Key R&D Program, Pudong Health and Family Planning Commission

**Research in context:** *Evidence before this study:* Several prognostic models for predicting mortality risk, progression to severe disease, or length of hospital stay in COVID-19 have been published.^1^ Commonly reported predictors of severe prognosis in patients with COVID-19 include age, sex, computed tomography scan features, C-reactive protein (CRP), lactic dehydrogenase, and lymphocyte count. Symptoms (notably dyspnoea) and comorbidities (e.g. chronic lung disease, cardiovascular disease and hypertension) are also reported to have associations with poor prognosis.^2^ However, most studies have not described the study population or intended use of prediction models, and external validation is rare and to date done using datasets originating from different Wuhan hospitals.^3^ Given different patterns of testing and organisation of healthcare pathways, external validation in datasets from other countries is required.

*Added value of this study:* This study used data from Wuhan, China to derive and internally validate multivariable models to predict poor outcome and death in COVID-19 patients after hospital admission, with external validation using data from King’s College Hospital, London, UK. Mortality and poor outcome occurred in 4.3% and 9.7% of patients in Wuhan, compared to 34.1% and 42.9% of patients in London. Models based on age, sex and simple routinely available laboratory tests (lymphocyte count, neutrophil count, platelet count, CRP and creatinine) had good discrimination and calibration in internal validation, but performed only moderately well in external validation. Models based on age, sex, symptoms and comorbidity were adequate in internal validation for poor outcome (ICU admission or death) but had poor performance for death alone.

*Implications of all the available evidence:* This study and others find that relatively simple risk prediction models using demographic, clinical and laboratory data perform well in internal validation but at best moderately in external validation, either because derivation and external validation populations are small (Xie et al^3^) and/or because they vary greatly in casemix and severity (our study). There are three decision points where risk prediction may be most useful: (1) deciding who to test; (2) deciding which patients in the community are at high-risk of poor outcomes; and (3) identifying patients at high-risk at the point of hospital admission. Larger studies focusing on particular decision points, with rapid external validation in multiple datasets are needed. A key gap is risk prediction tools for use in community triage (decisions to admit, or to keep at home with varying intensities of follow-up including telemonitoring) or in low income settings where laboratory tests may not be routinely available at the point of decision-making. This requires systematic data collection in community and low-income settings to derive and evaluate appropriate models.

## Introduction

Severe acute respiratory syndrome coronavirus-2 (SARS-CoV-2) is the novel coronavirus that causes coronavirus disease 2019 (COVID-19). A local outbreak of COVID-19 was first detected in Wuhan, China and rapidly spread to cause a pandemic.^4^

A key feature of the pandemic in all areas with large local epidemics is that healthcare systems are often rapidly overwhelmed. Effective and safe triage to ensure that those at highest risk of severe disease are appropriately escalated, and those at lower risk safely monitored, is critical to maintain healthcare system capacity for as long as possible. The same risk stratification is also needed to support targeted recruitment to both mechanistic and interventional research studies, since developing effective treatments of early disease is a key objective alongside vaccine development. Although risk stratification is important in all health systems, exactly how risk stratification is done will vary by country and by healthcare system. Across all systems though, important care pathway decision points include (1) who to test (particularly important when tests are in short supply); (2) who to hospitalize, and who to care for in the community (with or without telemonitoring or other proactive follow-up), and; (3) who to direct down different care pathways after admission. Since the severity of disease and risk of poor outcome varies by point on the pathway, there is a need for a range of risk stratification tools based on the data available at different stages of the care pathway (Figure 1), and/or in different healthcare systems.

**Figure 1:**
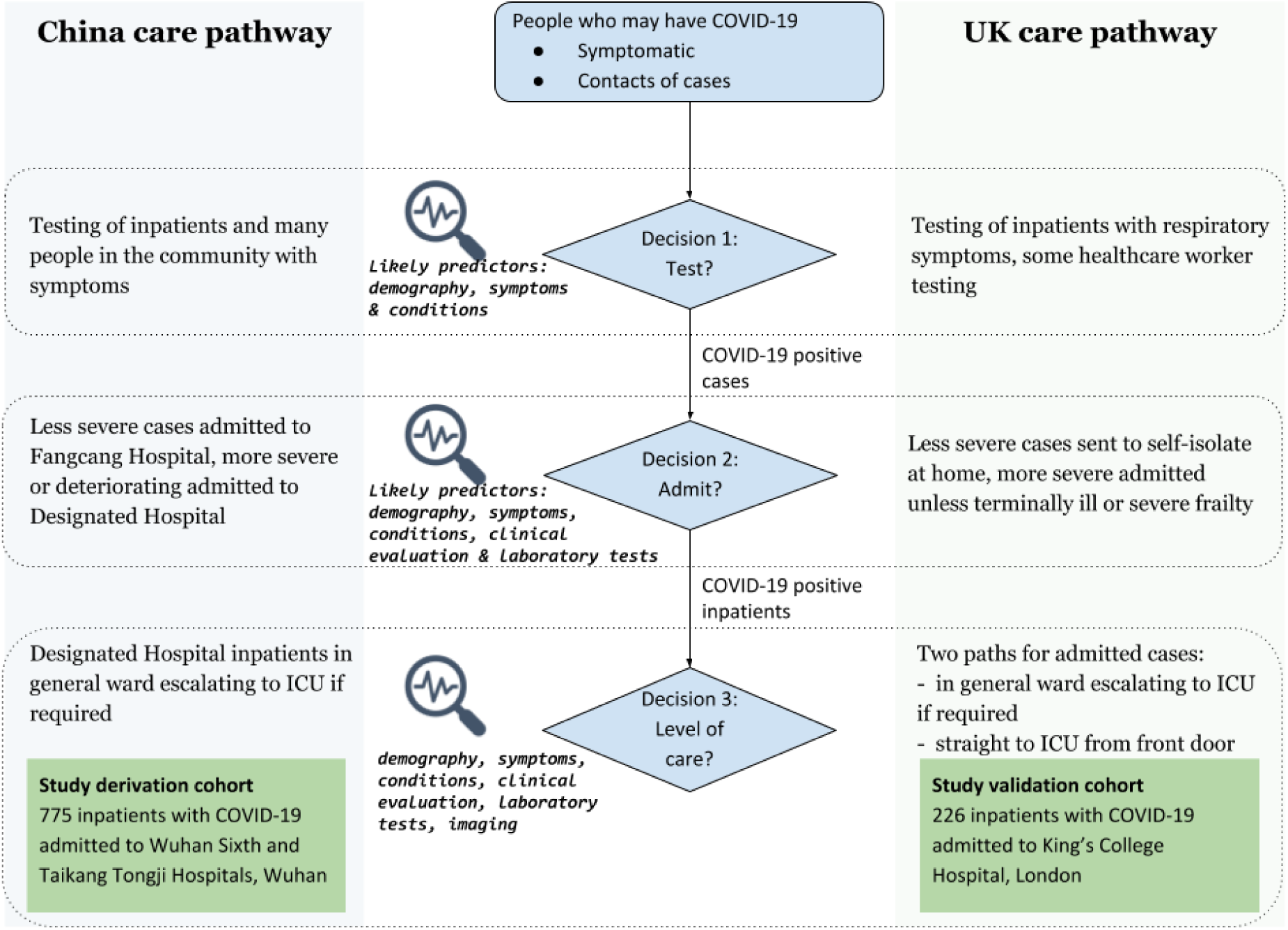
Comparison of COVID-19 patients care pathway in China and UK

A number of studies have identified patient characteristics associated with poor outcomes in COVID-19 using univariate analysis. Older age, being male, dyspnea and comorbidities (e.g. chronic obstructive airways disease [COPD], cardiovascular disease, hypertension) are associated with intensive care unit (ICU) admission,^2,5–11^ and comorbidities are also reported to be associated with an increased risk of death.^12,13^ However, many studies do not report adjusted associations and few examine laboratory findings, although there are reported univariate associations between the development of acute respiratory distress syndrome (ARDS) and a range of predictors including high fever (≥39°C), older age, and laboratory measures including neutrophilia, d-dimer and other evidence of coagulation and organ dysfunction.^14^ However, univariate analysis is usually a poor guide to risk because of confounding, most obviously between age and physical morbidities.^15^

Two studies have published multivariable/adjusted findings. Zhou et al examined risk factors for in-hospital death in 191 patients, finding significant adjusted associations with older age, higher Sequential Organ Failure Assessment (SOFA) score, lower lymphocyte count and increased d-dimer.^16^ Xie et al is the only published study explicitly aiming to create a risk prediction model for inpatient mortality for clinical use, with derivation in 299 patients admitted to one Wuhan hospital and external validation in 145 patients admitted to another Wuhan hospital.^3^ Mortality was very high in both derivation (51.8%) and validation (47.6%) cohorts. Factors associated with inpatient mortality were age, lower lymphocyte count, higher lactate dehydrogenase and lower peripheral capillary oxygen saturation (SpO2), and external validation showed reasonable performance (calibration was excellent but only after model recalibration to the validation dataset which will rarely be possible in clinical use).^3^ Neither study reported clinically relevant performance measures such as sensitivity, specificity and positive/negative predictive value.

The aim of this study was to derive and internally validate risk prediction models for poor outcome and death in a cohort of 775 inpatients with COVID-19 in Wuhan, China, with external validation in a cohort of 226 inpatients with COVID-19 in London, UK.

## Methods

### Methods are briefly described here and in detail in the supplementary file

#### Study design and participants

The multivariable risk prediction model was derived in a cohort of 775 adults with COVID-19 confirmed by RT-PCR admitted to one of two hospitals in Wuhan, China (Wuhan Sixth Hospital and Taikang Tongji Hospital). The model was externally validated in a cohort of 226 adult inpatients with confirmed COVID-19 in King’s College Hospital (KCH) NHS Foundation Trust, London, UK.

#### Statistical modelling

A logistic regression model was chosen for its good clinical interpretability. Models were fitted with L1-regularization (LASSO) to minimize overfitting. Two clinically relevant outcomes were defined: death and poor outcome (defined as: developing ARDS, receiving intubation or extracorporeal membrane oxygenation (ECMO) treatment, ICU admission and death). Predictor variables were participants’ demographic characteristics, premorbid conditions, symptoms and laboratory test results (around admission) as predictors. Combinations of predictors were made with consideration of information availability at points of the health care pathway (Figure 1):

1. Community triage: <u>D</u>emographic + premorbid <u>C</u>onditions + <u>S</u>ymptoms (DCS)
2. Hospital admission (full model): DCS + <u>L</u>aboratory results (DCSL)
3. Hospital admission (simpler model): Demographic + Laboratory results (DL)

Models are referred to as predictor-outcome (e.g. DCS-Death). For each model, predictors with coefficients significantly different to zero in cross-validation were selected. Optimal L1-regularization strength for preferred models was chosen by best C-index in cross-validation. All models were internally validated with cross-validation to select variables based on model fit and not just individual predictor statistical significance, and model performance metrics in the held-out partition were reported. To explore prediction cut-offs to use to inform clinical decision-making, we grouped patients into low-, high- and very high-risk groups based on predicted probabilities obtained from the final models. Cut-offs to define the low and high-risk groups were arbitrarily assigned to be approximately 10-fold lower and 8-fold higher than the outcome percentage in the whole dataset. Models using DL predictors were externally validated in the KCH cohort, using two outcomes: death and poor outcome (defined as ICU admission or death).

#### Ethics and governance

The derivation study was approved by the Research Ethics Committee of Shanghai Dongfang Hospital and Taikang Tongji Hospital. The external validation study operated under London South East Research Ethics Committee (reference 18/LO/2048) approval granted to the King’s Electronic Records Research Interface (KERRI).

## Results

### Description of the patient cohort

The derivation cohort consisted of 775 adult in-patients with COVID-19 in Wuhan (Table 1). The median age was 61 years (IQR 50-68, range 18-96), 48.9% were men, and 345 (44.5%) had comorbidity (most commonly hypertension (30.8%), diabetes mellitus (13.7%) and heart disease (11.0%)). The most common symptoms on admission were fever (68.6%) and cough (68.1%), followed by fatigue (54.3%) and dyspnoea (45.8%). Median lymphocyte count was 1.4 (IQR 1.1-1.9) 10^9/L, neutrophil count 3.5 (2.7-4.9) 10^9/L, platelet count 224 (180-274) 10^9/L, CRP 2.7 (0.9-14.1) mg/L and creatinine 63.9 (54.1-76.4) μmol/L.

**Table 1:** Characteristics of the derivation and external validation cohorts

|  | Wuhan cohort<br>(n=775) |  | KCH cohort<br>(n=226) |  |
| --- | --- | --- | --- | --- |
| **Demographic** |  | No. (%) |  | No. (%) |
| Age (median (IQR)) | 775 | 61 (50-68) | 226 | 74 (59-85) |
| Sex (male) | 775 | 379 (48.9%) | 226 | 124 (54.9%) |
| **Outcome** |  |  |  |  |
| Death | 773 | 33 (4.3%) | 226 | 77 (34.1%) |
| Poor outcome | 772 | 75 (9.7%) | 226 | 97 (42.9%) |
| **Premorbid Conditions** |  |  |  |  |
| Chronic lung disease | 771 | 48 (6.2%) | .. | .. |
| Diabetes Mellitus | 771 | 106 (13.7%) | .. | .. |
| Immunocompromised | 771 | 12 (1.5%) | .. | .. |
| Malignancy | 771 | 24 (3.1%) | .. | .. |
| Hypertension | 771 | 239 (30.8%) | .. | .. |
| Heart disease | 771 | 85 (11.0%) | .. | .. |
| Chronic renal disease | 771 | 25 (3.2%) | .. | .. |
| **Symptoms** |  |  |  |  |
| Fever | 773 | 532 (68.6%) | .. | .. |
| Cough | 773 | 528 (68.1%) | .. | .. |
| Fatigue | 773 | 421 (54.3%) | .. | .. |
| Dyspnoea | 773 | 355 (45.8%) | .. | .. |
| Diarrhoea | 773 | 36 (4.6%) | .. | .. |
| **Laboratory tests (median (IQR))** |  |  |  |  |
| Neutrophil count (10<sup>9</sup>/L) | 739 | 3.5 (2.7-4.9) | 226 | 4.9 (3.4-6.9) |
| Lymphocyte (10<sup>9</sup>/L) | 743 | 1.4 (1.1-1.9) | 226 | 1.1 (0.7-1.6) |
| Platelet (10<sup>9</sup>/L) | 732 | 224 (180-274) | 226 | 211 (160-284) |
| C-reactive protein (mg/L) | 684 | 2.7 (0.9-14.1) | 226 | 56.0 (21.1-115.5) |
| D-dimer (mg/L) | 581 | 0.4 (0.2-1.1) | .. | .. |
| Creatinine (μmol/L) | 728 | 63.9 (54.1-76.4) | 226 | 90.0 (68.0-126.0) |
| Alanine aminotransferase (IU/L) | 723 | 22.9 (14.8-39.5) | .. | .. |

Data are median (IQR) or number (%). Two patients had missing data for death and three patients had missing data for poor outcome

The validation cohort consisted of 226 adult patients admitted to Kings College Hospital, London with a positive COVID-19 RT-PCR, and with complete data for the laboratory tests included in the derivation risk prediction model (Table 1). Median age was 74 years (IQR 59-85, range 19-98) and 54.9% were men. Data on symptoms and comorbidity were not extracted. Median lymphocyte count was 1.1 (IQR 0.7-1.6) 10^9/L, neutrophil count 4.9 (3.4-6.9) 10^9/L, platelet count 211 (160-284) 10^9/L, CRP 56.0 (21.1-115.5) mg/L, and creatinine 90.0 (68.0-126.0 μmol/L.

Median lymphocyte count and platelet count was lower, and median age, proportion male, neutrophil count, CRP and creatinine higher in the KCH validation compared to the Wuhan derivation cohort. In the 7 derivation cohort, 33 (4.3%) died and 75 (9.7%) had pre-defined poor outcome (ARDS, intubation, ECMO, ICU admission or death) (two patients had missing data for death and three patients had missing data for poor outcome). Both outcomes were much more common in the validation cohort where 77 (34.1%) died, and 97 (42.9%) of patients experienced a poor outcome (ICU admission or death).

Univariate associations of patient characteristics and outcomes are summarised in Table S1, with findings broadly consistent with previously reported univariate associations.^2,12,13^ The strongest observed univariate associations were increased risk of each outcome with increasing age, the presence of prior comorbidity (particularly chronic lung disease, heart disease, immunocompromise, chronic renal disease and hypertension), the presence of dyspnoea, and laboratory values (increased risk with higher neutrophil count, CRP and creatinine, and with lower lymphocyte count and platelet count).

### Derivation of prediction models in the Wuhan cohort

Table 2 summarises multivariable odds ratios of models for both outcomes (Table S2 reports the predictor coefficients with more precision), and Table 3 details model performance in internal and external validation. In adjusted analysis, underlying conditions and symptoms were no longer associated with either outcome with the exception of dyspnoea which was associate with poor outcome, although confidence intervals are wide meaning that a type 2 error cannot be ruled out.

**Table 2:** Multivariable logistic regression analysis of risk factors associated with Death and Poor outcome in COVID-19 patients in the derivation cohort

|  | **Death**<br>**Odds Ratio† (95%CI)** |  |  | **Poor outcome**<br>**Odds Ratio (95%CI)** |  |  |
| --- | --- | --- | --- | --- | --- | --- |
| Predictor | DCS* 30/769 (3.9%) | DCSL 19/651 (2.9%) | DL 20/653 (3.1%) | DCS 72/768 (9.4%) | DCSL 57/651 (8.8%) | DL 58/653 (8.9%) |
| **Demographic** |  |  |  |  |  |  |
| Age | 1.05 (1.01-1.08) | 1.00 (0.95-1.05) | 1.01 (0.96-1.07) | 1.04 (1.02-1.07) | 1.02 (0.99-1.05) | 1.03 (1.000-1.058) |
| Sex (male) | 1.37 (0.62-3.03) | 0.42 (0.12-1.49) | 0.45 (0.13-1.50) | 1.23 (0.72-2.09) | 0.60 (0.28-1.29) | 0.62 (0.30-1.28) |
| **Premorbid Conditions** |  |  |  |  |  |  |
| Chronic lung disease | 3.00 (1.07-8.37) | 1.50 (0.26-8.52) | .. | 3.42 (1.61-7.25) | 2.63 (0.81-8.51) | .. |
| Diabetes Mellitus | 1.56 (0.60-4.09) | 1.37 (0.35-5.38) | .. | 1.00 (0.50-2.01) | 0.75 (0.28-2.01) | .. |
| Immunocompromised | 3.97 (0.64-24.65) | .. | .. | 4.19 (1.05-16.75) | 4.82 (0.63-36.76) | .. |
| Malignancy | 1.02 (0.15-7.05) | 1.00 (0.13-7.73) | .. | 1.00 (0.25-3.99) | 0.49 (0.08-2.88) | .. |
| Hypertension | 1.41 (0.62-3.23) | .. | .. | 1.94 (1.11-3.39) | 1.62 (0.73-3.60) | .. |
| Heart disease | 1.97 (0.77-5.02) | .. | .. | 2.10 (1.08-4.08) | 2.09 (0.80-5.43) | .. |
| Chronic renal disease | 3.75 (0.92-15.30) | .. | .. | 2.70 (0.90-8.06) | 0.70 (0.11-4.62) | .. |
| **Symptoms** |  |  |  |  |  |  |
| Fever | .. | .. | .. | 1.03 (0.57-1.85) | 1.18 (0.53-2.66) | .. |
| Cough | 2.10 (0.70-6.32) | 2.57 (0.53-12.47) | .. | 1.00 (0.53-1.88) | 0.55 (0.25-1.22) | .. |
| Fatigue | .. | .. | .. | 1.10 (0.62-1.96) | 1.22 (0.56-2.66) | .. |
| Dyspnoea | 4.59 (1.79-11.79) | 2.35 (0.65-8.47) | .. | 2.97 (1.64-5.39) | 3.47 (1.55-7.75) | .. |
| Diarrhoea | 1.00 (0.11-8.94) | .. | .. | 0.81 (0.17-3.78) | 0.07 (0.00-19.37) | .. |
| **Laboratory tests** |  |  |  |  |  |  |
| Neutrophil count (10<sup>9</sup>/L) | .. | 1.20 (1.03-1.39) | 1.25 (1.09-1.44) | .. | 1.28 (1.12-1.47) | 1.29 (1.14-1.46) |
| Lymphocyte count (10<sup>9</sup>/L) | .. | 0.24 (0.05-1.04) | 0.27 (0.06-1.15) | .. | 0.29 (0.12-0.72) | 0.31 (0.14-0.68) |
| Platelet count (10<sup>9</sup>/L) | .. | 0.99 (0.987-1.001) | 0.99 (0.988-1.002) | .. | 1.00 (0.992-1.001) | 1.00 (0.993-1.001) |
| C-reactive protein (mg/L) | .. | 1.01 (1.003-1.025) | 1.01 (1.004-1.024) | .. | 1.01 (0.998-1.018) | 1.01 (1.000-1.017) |
| Creatinine (μmol/L) | .. | 1.00 (0.998-1.005) | 1.00 (0.998-1.005) | .. | 1.01 (1.005-1.021) | 1.01 (1.003-1.016) |

\* Initials specifying predictor categories examined in each model: D – Demographic, C - Premorbid Conditions, S – Symptoms, L - Laboratory Results. Data in the table head are number with outcome/number with complete data included in model (% with outcome). Two patients had missing data for death and three patients had missing data for poor outcome.
† For continuous value (age and laboratory tests), the OR interpretation is that the odds of having the outcome increase a value of OR-1 for a one-unit increase in the value. For example, for Death DCS model, the OR for Death is 1.06. This means that every 1 year increase in age is associated with a 6% increase in the odds of death (e.g. across the 18 year interquartile range of age, this equates to an OR of 2.85).
.. indicates the corresponding predictor is not included in the model

For the outcome of death, DCS-Death, DCSL-Death and DL-Death models achieved C-indices of 0.79, 0.89 and 0.91, respectively (Table 3 and Figure S1A). Calibration evaluated using calibration-in-the-large and calibration slope was poor for DCS-Death and DCSL-Death models, but good for DL-Death (calibration-in-the-large 0.19 [perfect calibration=0], calibration slope 0.98 [perfect calibration=1]).

**Table 3:** Internally and externally validated performances of prediction models: discrimination, calibration and clinical usefulness

|  | Internal cross-validation |  |  |  |  |  | Externally validation |  |
| --- | --- | --- | --- | --- | --- | --- | --- | --- |
|  | Death |  |  | Poor outcome |  |  | Death | Poor outcome |
|  | DCS<sup>a</sup><br>N: 769 (3·9%) | DCSL<sup>a</sup><br>N: 651 (2·9%) | DL<sup>a</sup><br>N: 653 (3·1%) | DCS<sup>a</sup><br>N: 768 (9·4%) | DCSL<sup>a</sup><br>N: 651 (8·8%) | DL<sup>a</sup><br>N: 653 (8·9%) | DL<sup>a</sup><br>N: 226 (34·1%) | DL<sup>a</sup><br>N: 226 (42·9%) |
| **Discrimination and calibration** |  |  |  |  |  |  |  |  |
| C-index | 0·79 | 0·89 | 0·91 | 0·80 | 0·88 | 0·88 | 0·74 | 0·72 |
| Calibration in the large<br>(95% CI)<sup>b</sup> | 0·384<br>(-0·077-0·846) | 0·316<br>(-0·078-0·711) | 0·189<br>(-0·104-0·481) | 0·132<br>(-0·133-0·397) | 0·059<br>(-0·169-0·287) | 0·008<br>(-0·131-0·147) | 0·251<br>(0·051-0·451) | 0·276<br>(0·086-0·466) |
| Calibration slope<br>(95% CI)<sup>b</sup> | 0·287<br>(-0·544-1·117) | 0·460<br>(-0·288-1·208) | 0·981<br>(0·244-1·718) | 0·855<br>(0·340-1·370) | 0·894<br>(0·507-1·281) | 1·036<br>(0·784-1·288) | 0·543<br>(0·202-0·885) | 0·540<br>(0·212-0·868) |
| **Clinical Usefulness<sup>c</sup>** |  |  |  |  |  |  |  |  |
| Positive predictive value | 0·01 | 0·42 | 0·52 | 0·53 | 0·64 | 0·72 | 0·69 | 0·67 |
| Negative predictive value | 0·96 | 0·98 | 0·98 | 0·91 | 0·94 | 0·94 | 0·71 | 0·65 |
| Sensitivity | 0·003 | 0·25 | 0·33 | 0·09 | 0·37 | 0·36 | 0·23 | 0·40 |
| Specificity | 0·997 | 0·99 | 0·99 | 0·99 | 0·98 | 0·99 | 0·95 | 0·85 |
| **Clinical Usefulness<sup>d</sup>**<br>**Low risk vs high/very high** |  |  |  |  |  |  |  |  |
| Positive predictive value | 0·07 | 0·25 | 0·25 | 0·19 | 0·22 | 0·17 | 0·55 | 0·44 |
| Negative predictive value | 0·997 | 0·998 | 0·997 | 0·99 | 0·99 | 0·99 | 0·85 | 0·74 |
| Sensitivity | 0·97 | 0·95 | 0·90 | 0·96 | 0·95 | 0·97 | 0·77 | 0·95 |
| Specificity | 0·51 | 0·92 | 0·91 | 0·57 | 0·67 | 0·54 | 0·68 | 0·11 |
| **Clinical Usefulness<sup>d</sup>**<br>**Low/high risk vs very high** |  |  |  |  |  |  |  |  |
| Positive predictive value | 0·32 | 0·32 | 0·28 | 0·67 | 0·60 | 0·60 | 0·58 | 0·72 |
| Negative predictive value | 0·97 | 0·995 | 0·99 | 0·93 | 0·96 | 0·96 | 0·78 | 0·65 |
| Sensitivity | 0·30 | 0·84 | 0·75 | 0·25 | 0·53 | 0·53 | 0·58 | 0·61 |
| Specificity | 0·97 | 0·95 | 0·94 | 0·99 | 0·97 | 0·96 | 0·78 | 0·75 |

In the DL-Death model, increased blood neutrophil counts (OR 1.25 [all OR for continuous variables are per one unit increase in predictor values], p=0.002) and CRP levels (OR 1.01, p=0.008) were significantly associated with death. Other variables were not statistically significantly associated with death but included because they improved model fit.

Using a probability of 0.5 as the prediction cut-off for the DL-Death model, sensitivity was 0.33, positive predictive value 0.52, specificity 0.99, and negative predictive value 0.98, which is not adequate for clinical use. Using the two alternative probability cut-offs to define low, high, and very high risk groups, the percentage who died in the low, high, and very high risk groups were 0.34% (n=2/580), 15.0% (n=3/20) and 28.3% (n=15/53), respectively (Figure 2A-B, Table S3). Notably, the model defined a low-risk group comprising 88.8% of patients with very high negative predictive value (0.997) (Table 3) as well as a very-high risk group comprising 8.1% of patients in whom 75% of observed deaths occurred.

**Figure 2:**
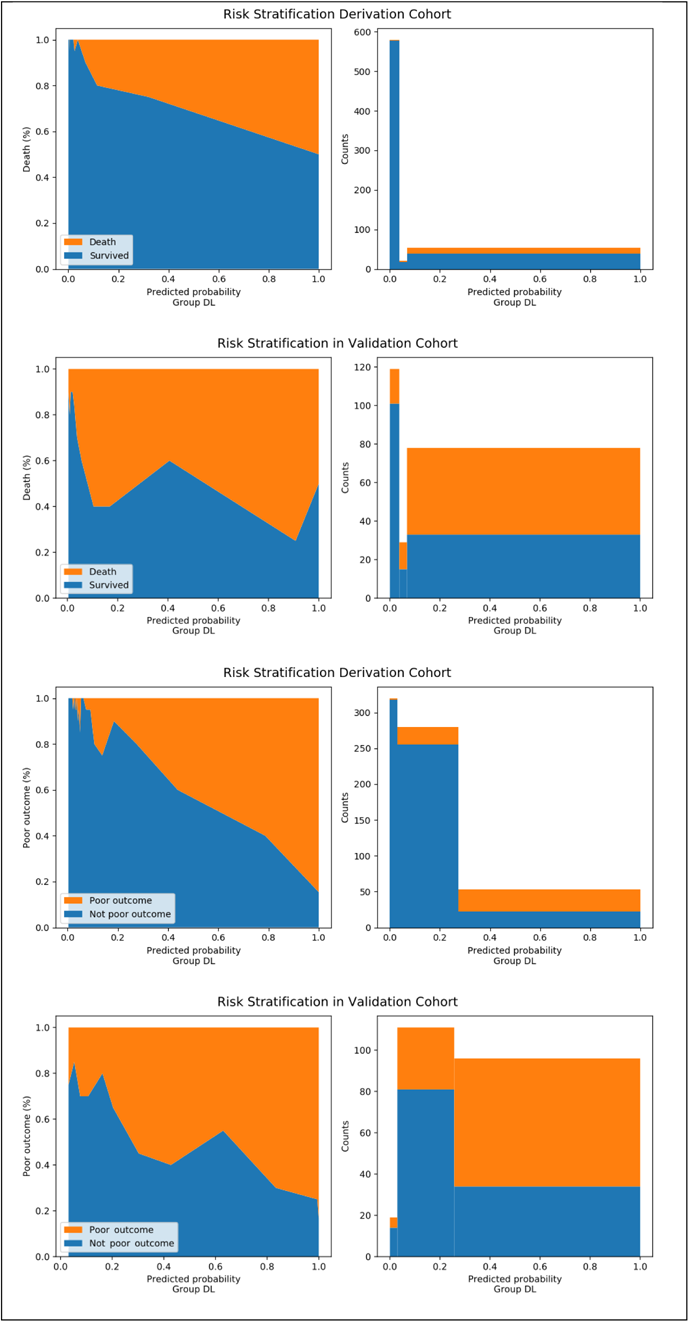
Risk stratification of COVID-19 patients using predicted probability of DL-Death (top two panels) and DL-Poor models (bottom two panels)

For poor outcome, the DCS-Poor, DCSL-Poor and DL-Poor models achieved C-indices of 0.80, 0.88 and 0.88, respectively (Table 3, Figure S1B). Calibration was better than for models predicting death, and was very good for the DL-Poor model (calibration-in-the-large 0.01, calibration slope 1.04).

In the DL-Poor model, risk of poor outcome of COVID-19 was associated with increased blood neutrophil counts (OR 1.29 [all OR for continuous variables are per one unit increase in predictor value], p=<0.001), increased creatinine levels (OR 1.01, p=0.002) and decreased blood lymphocyte counts (OR 0.31, p=0.004). Age (OR 1.03, p=0.053) and CRP (OR 1.01, p=0.060) were strongly associated but marginally not statistically significant in adjusted models. Other variables were not statistically significantly associated with death but included because they improved model fit.

Using a probability of 0.5 as the prediction cut-off for the DL-Poor model, sensitivity was 0.36, positive predictive value 0.72, specificity 0.99, and negative predictive value 0.94, which is not ideal for clinical use. Using the two alternative probability cut-offs, the percentage of patients with poor outcome in the low, high, and very high-risk groups were 0.63% (n=2/320), 8.9% (n=25/280) and 58.5% (n=31/53), respectively (Figure 2E-F). Notably, the model defined a low-risk group comprising 49.0% of patients with very high negative predictive value (0.99) (Table 3) as well as a very-high risk group comprising 8.1% of patients with high positive predictive value (0.60) in which 53% of observed poor outcomes occurred.

### External validation of the model in KCH cohort

Table 3 shows the performance (discrimination, calibration and clinical usefulness) of the DL-Death and DL-Poor models without any re-calibration. Both models showed good discrimination measured by the C-index (death: 0.74; poor outcome: 0.72), but calibration was only fair with calibration-in-the-large 0.25 for death and 0.28 for poor outcome, and calibration slope 0.54 for death and 0.54 for poor outcome. Calibration plots are shown in Figure S2, and show some under-prediction at low levels of risk. Using the default probability cut-off of 0.5, sensitivity was poor (0.23 for death; 0.40 for poor outcome), specificity good to excellent (0.95; 0.85), PPV (0.69; 0.67) and NPV (0.71; 0.65) were fair, given outcome prevalence.

Using the derivation model probability cut-offs for low, high and very high risk groups, the DL-Death model categorized the external validation cohort into groups with death rates of 16.5% (n=20/121), 42.9% (n=12/28) and 58.4% (n=45/77). Sensitivity at the low-high risk cut-off was 0.77, specificity 0.68, PPV 0.55 and NPV 0.85, and at the high-very high risk cut-off sensitivity 0.58, specificity 0.78, PPV 0.58 and NPV 0.78.

Using the derivation model probability cut-offs for low, high and very high risk groups, the DL-Poor model categorized the KCH cohort into groups with outcome rates of 26.3% (n=5/19), 28.4% (n=33/116) and 64.8% (n=59/91). Sensitivity at the low-high risk cut-off was 0.95, specificity 0.11, PPV 0.44 and NPV 0.74, and at the high-very high risk cut-off sensitivity 0.61, specificity 0.75, PPV 0.72 and NPV 0.65.

## Discussion

We derived and internally validated models to predict the risk of death and poor outcome for COVID-19 patients using a cohort of inpatients in Wuhan, China with a relatively low risk of death (4.3%) and poor outcome (9.7%). Four groups of predictors were examined: demographic information, premorbid conditions, symptoms and laboratory results. Models including demographic and laboratory data (DL models) had the best performance, with older age, higher neutrophil count, lower lymphocyte count, lower platelet count, higher CRP and higher creatinine being predictive of death and poor outcome. In internal validation, DL model performance was good, and it was possible to define low, high and very-high risk groups for both outcomes. DL models were externally validated in a cohort of inpatients in London, UK with eight-fold higher risk of death (34.1%) and four-fold higher risk of poor outcome (42.9%) reflecting different care pathways in the two countries (Figure 1) and therefore different casemix of those admitted (Table 1). In this very different cohort, DL model performance was only fair to moderate in the external validation dataset, and further external validation is required in datasets with different casemix and severity.

Our study has several strengths. First, model derivation used a relatively large cohort of inpatients with COVID-19 from Wuhan, China with 99.6% ascertainment of endpoints, and predictors measured on admission. Derivation and internal validation of prediction models was robust. Second, we externally validated parsimonious risk prediction models using a dataset from a different country with a very different pattern of COVID-19 admission (London, UK). Third, the study is reported in accordance with the TRIPOD guidelines^17^ including detailed methods reporting in the supplementary file to facilitate reproducibility and further external validation.^1^

The study also has a number of limitations. First, the clinical datasets were collected when healthcare services were under severe strain. Data extraction sought to ensure consistency and accuracy, but was not blind to outcome, and there is missing data in both datasets. Second, the datasets used are smaller than ideal (although as large as or larger than previous studies), and there are relatively few deaths in particular. Our analytical approach aimed to minimise overfitting, but further research using larger, federated datasets is clearly required. Third, clinical assessments at admission such as SpO2 are likely to be important predictors of short-term outcome,^3^ but were not available in either dataset. Fourth, our external validation dataset has very different case-mix where spectrum effects are likely to contribute to lower prediction model performance, and only has follow-up to a fixed date (period range: [6-39] days, although this is a reasonable time-horizon to inform clinical decision-making at hospital admission). Finally, all data available is for people with PCR-diagnosed COVID-19 who are admitted to hospital (decision 3 in Figure 1). Although the Wuhan cohort includes many people with less severe disease, in the validation cohort most admitted patients are likely to have severe disease. The findings therefore cannot be assumed to be applicable to decisions made earlier in the course of disease (decision 2 in Figure 1).

Our univariate findings are similar to other studies reporting univariate associations. Older patients and those with dyspnoea or premorbid conditions (e.g. chronic lung disease, hypertension, cardiovascular disease) are consistently reported to be more susceptible to in-hospital death or poor outcome of COVID-19.^2,6,8,9,12,13,18^ Lymphocytopenia, neutrophilia, and markers of inflammation (e.g. CRP), cell death (e.g. lactate dehydrogenase (LDH)) and abnormal coagulation (e.g. d-dimer) have all also been reported as associated with poor outcome.^14^ However, the majority of these studies are small and only reported univariate associations.

Our findings are similar to the few studies reporting multivariable associations, although the predictors included are not identical.^3,16^ Age is a very strong predictor of outcome, and accounting for age reduces associations with comorbidity and symptoms (although no study including our own is large enough to rule out significant type 2 error i.e. that these are truly *not* independently associated with outcomes). Lymphocyte count is included in all three models, but Zhou et al also included d-dimer and SOFA score, whereas Xie et al included LDH and SpO2, compared to our inclusion of neutrophil count and CRP. The three models are therefore based on demography, (different) laboratory measures, and (different) clinical measures. The choice of predictors in all three studies is driven by data availability, and by initial univariate analysis identifying different features to model (SOFA score for example is a good predictor in Zhou et al,^16^ but not in Xie et al’s study.^3^ There is a need for larger studies to derive and validate more optimal models suitable for use at different points in the natural history of COVID-19, since for example, comorbidity may (or may not) be an important predictor of admission but less important than laboratory markers for predicting prognosis in those admitted.

Xie et al’s prediction model was externally validated in a similar dataset from Wuhan with very high mortality in both datasets (derivation cohort 51.8% died, validation cohort 47.6%). Discrimination in external validation was excellent, but calibration was poor until the model intercept and slope were recalibrated to the external validation dataset. We chose not to recalibrate our model to the external dataset, since this will hardly ever be feasible in a tool intended for clinical use for an epidemic disease (and Xie et al’s nomogram intended for use by clinicians is also based on the derivation model). Xie et al did not publish clinically relevant performance metrics such as sensitivity, specificity, PPV and NPV, making it difficult to evaluate clinical utility.

The findings of this and previous studies are consistent with demography and routinely available laboratory measurements being useful markers of prognosis at the time of hospital admission, indicating that simple risk prediction tools may usefully inform clinical decision-making at this point in the care pathway. However, COVID-19 risk prediction models need external validation before widespread clinical use, because severity and prognosis depends on the setting and system of care. Our Wuhan derivation cohort is closer to a general population cohort (mortality 4.3%) than either our validation cohort in London (mortality 34.1%) or the Wuhan cohorts in other studies (mortality 28.3%,^16^ and 51.8% derivation and 47.6% validation^3^). It may therefore be better suited to decisions about who to admit and who can safely be sent home to self-isolate (decision 2 in Figure 1) than to identifying those at highest risk of poor outcome and death in people with more severe disease, for example at the point of potential ICU admission. All proposed prediction models therefore require repeated external validation in multiple datasets to identify the contexts in which they can be most effectively deployed. The DCS and DL models are available as prediction tools in a web portal but we emphasise that they should be used with caution, particularly in settings with different rates of poor outcomes, and are not a replacement for clinical judgement (https://covid.datahelps.life/prediction/).

Although very challenging under epidemic conditions, systematic data collection during clinical care and rapid data analysis is therefore essential to understand the natural history of COVID-19 to inform clinical decision-making along the entire care pathway (Figure 1). Of note is that our models based solely on demography, symptoms and premorbid conditions did not perform optimally in internal validation, which may indicate that community assessment would optimally include simple blood tests. Further research is needed to examine this, and the relative value of laboratory tests compared to clinical examination (e.g. pulse, respiratory rate, SpO2). Key gaps are in data collection and analysis in early disease and in the community (since minimising admission when it is safe to do so is critical for keeping hospital services functioning) and at the point of ICU admission (since there may or may not be populations where mechanical ventilation is predictably futile).

The observed associations between raised neutrophil^19^ and low lymphocyte counts^20^ with outcomes highlights that understanding and delineating the innate and adaptive immune cascade may inform potential interventional strategies. These include targeting viral replication alongside directed anti-inflammatory agents to mitigate the inflammatory cascade.^21^ The timing of such interventions is likely to be crucial, and risk prediction models therefore have the additional potential to support targeted and timely treatment initiation.

## Conclusion

Risk prediction tools have considerable potential to inform clinical decision-making at different points in the COVID-19 care pathway. Existing models, including our own, perform well in derivation datasets but robust and repeated external validation is required before widespread clinical use. Collaborative research to derive and validate tools using larger datasets from all points on the clinical pathway is urgently needed.

## Data Availability

Data from patient records used in the study will not be available due to inability to fully anonymise up to the Information Commissioner Office (ICO) standards and ethical requirements. A subset of the dataset limited to anonymisable information is available on request to researchers with suitable training in information governance and human confidentiality protocols subject to approval by the King's College Hospital Information Governance committee and Shanghai East Hospital committee; applications for research access to KCH cohort data should be sent to and applications for research access to Wuhan cohort data should be sent to. This dataset cannot be released publicly due to the risk of re-identification of such granular individual-level data. Risk prediction models are published at (https://covid.datahelps.life/prediction/).

## Author contribution

QL, HW, BG, HZ, TS, and XW conceived the study. All data from the Wuhan cohort were extracted by XZ, JS, PZ, and CL, double-checked by KW and XW, with disagreements resolved by involving a third independent reviewer (QL). Data from the KCH cohort were extracted by DB and reviewed by JTT. HZ, TS, BG, KD and HW analysed the data, with HZ, TS, BG, KD, HW, XW, RD and JTT leading interpretation of findings. All authors participated in the preparation and approval of the Article.

## Acknowledgements

HW and HZ are supported by Medical Research Council and Health Data Research UK Grant (MR/S004149/1), Industrial Strategy Challenge Grant (MC_PC_18029) and Wellcome Institutional Translation Partnership Award (PIII054). RD is supported by the National Institute for Health Research (NIHR) Biomedical Research Centre at South London and Maudsley NHS Foundation Trust and King’s College London. DMB is funded by a UKRI Innovation Fellowship as part of Health Data Research UK MR/S00310X/1 (https://www.hdruk.ac.uk). KD is supported by LifeArc STOPCOVID award. This work uses data provided by patients and collected by the NHS as part of their care and support. XW is supported by National Natural Science Foundation of China (grant number:81700006). QL is supported by National Key R&D Program (2018YFC1313700), National Natural Science Foundation of China (grant number: 81870064) and the “Gaoyuan” project of Pudong Health and Family Planning Commission (PWYgy2018-06). The views expressed are those of the authors and not necessarily those of the NHS, the NIHR, or the Department of Health and Social Care. We acknowledge Lingyu Ran and Yongsheng Du for their contribution in data collection.

